# Clonal Hematopoiesis of Indeterminate Potential Status is Associated with Left Main Artery Stenosis

**DOI:** 10.1101/2023.02.10.23285708

**Authors:** J. Brett Heimlich, Michael A. Raddatz, John Wells, Caitlyn Vlasschaert, Sydney Olson, Marcus Threadcraft, Kristoff Foster, Emmanuel Boateng, Kelsey Umbarger, Yan Ru Su, Dan M. Roden, Colin M. Barker, Alexander G. Bick

## Abstract

Clonal hematopoiesis of indeterminate potential (CHIP) occurs as a result of acquired mutations in bone marrow progenitor cells. CHIP confers a twofold risk of atherosclerotic cardiovascular disease (ASCVD). However, there is limited data regarding specific cardiovascular phenotypes in this population. We recruited patients from the Vanderbilt University Medical Center cardiac catheterization laboratory and performed next generation sequencing to determine CHIP status. Multivariable logistic regression models and proportional odds models were used to assess the association between CHIP status and coronary angiography. We find nearly 1 in 5 patients undergoing coronary angiography have a CHIP mutation. Those with CHIP had a higher risk of having left main coronary artery disease compared to non-CHIP carriers. We additionally find that those with a specific CHIP mutation, ten eleven translocase 2 *(TET2)* has a larger effect size on left main stenosis compared with other CHIP mutations. This is the first description of a specific atherosclerotic phenotype in CHIP and serves as a basis for understanding enhanced morbidity and mortality in CHIP.

## Text

Clonal hematopoiesis of indeterminate potential (CHIP) occurs as a result of acquired mutations in bone marrow progenitor cells. CHIP confers a twofold risk of atherosclerotic cardiovascular disease (ASCVD) in large population studies, likely mediated through elevated inflammatory signaling^1^. Retrospective analysis of CHIP in patients from the CULPRIT-SHOCK trial revealed that among patients in cardiogenic shock after myocardial infarction, those with CHIP also suffered increased morbidity and mortality when compared to non-CHIP patients^2^. Despite these population-based findings, there are limited data detailing individual-level disease manifestations in humans. To date, there have been no prospective studies of CHIP in the cardiovascular setting, and the current literature lacks deep characterization of cardiovascular phenotypes, including coronary angiography.

We recruited patients from the Vanderbilt University Medical Center (VUMC) cardiac catheterization laboratory from May 2003 – April 2005 and August 2021 – August 2022 under the VUMC Institutional Review Board approval #090828. Adult patients without existing hematologic malignancy undergoing a planned cardiac procedure were enrolled. Next generation sequencing was performed on peripheral blood mononuclear cells using a custom sequencing panel and CHIP variant status determined as previously described^3^. Angiography results, laboratories, and anthropometric data were extracted from the electronic health record and compiled in a database during a single extraction period in December 2022. Study personnel were blinded to CHIP status until the final analysis was performed. Multivariable logistic regression models and proportional odds models were used to assess the association between CHIP status and binary and quantitative variables, respectively. Odds ratios in these models were adjusted by age, age^2^, sex, race, ethnicity, hyperlipidemia, statin use, hypertension, and diabetes status.

We sequenced 1,469 patients, and 1,149 had complete angiographic data. Of these, 214 (18.6%) had CHIP mutations with a variant allele frequency of >2% representing the expected spectrum of mutations **(Fig. 1A)**. CHIP patients were significantly older (68 vs. 59 years, p = 7.7 × 10^−16^). CHIP carrier status was significantly correlated with any stenosis and degree of stenosis in the left main (LM) coronary artery (OR 1.87 [1.28-2.74]; P = 0.0013 and OR 2.12 [1.42-3.15]; P = 2.2 × 10^−4^, **Fig. 1B**). This finding was not observed in any other coronary territory. This association was stronger among patients with Ten eleven translocase 2 *(TET2)* CHIP, both for any LM stenosis (OR 2.98 [1.48 – 5.96]; P = 0.0021) and the degree of LM stenosis (OR 3.52 [1.78 – 6.95]; P = 2.9 × 10^−4^). *TET2* CHIP was also correlated with obstructive (>70%) stenosis in any vessel (OR 2.28 [1.15 – 4.52]; P = 0.019), and the number of obstructive stenoses across coronary vessels (OR 1.83 [1.04 – 3.23]; P = 0.037).

**Figure 1.**
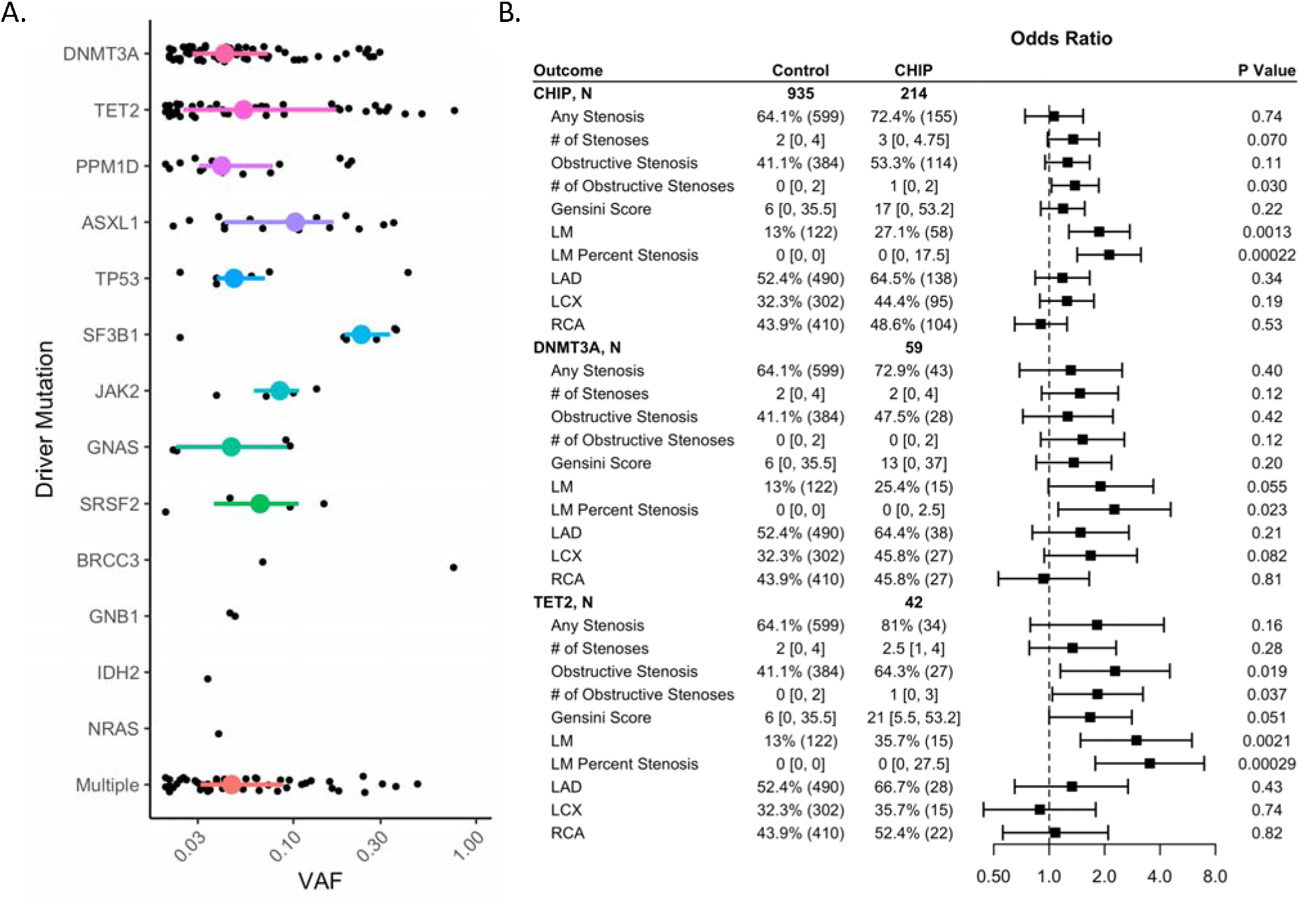
Variant allele fraction (VAF) distribution by driver gene (A) and adjusted odds ratios for the association of CHIP with coronary angiogram findings (B). Numeric data is displayed as percentage (N) for binary outcomes, and median [interquartile range] for continuous data (B). LM = left main; LAD = left anterior descending; LCX = left circumflex; RCA = right coronary artery.

CHIP is a known risk factor for the development ASCVD; however, there have been no cohort-based studies evaluating specific cardiovascular phenotypes, leaving a critical knowledge gap. In a single-center, prospective, observational cohort study of patients undergoing cardiac catheterization, we report nearly 1 in 5 had a detectable CHIP mutation. Remarkably, we find a significant association between CHIP status and left main artery stenosis by coronary angiography. A larger effect size for LM stenosis and an association for obstructive lesions in other vessels was seen for *TET2* CHIP, corroborating previous data that this subgroup exhibits more severe cardiovascular phenotypes, driven by inflammatory signaling^4^. We were unable to derive an effect of increasing clone size on any outcome due to limited data. Our findings reveal a population enriched for CHIP carriers in the cardiac catheterization laboratory, an important observation for future studies focused on secondary prevention in CHIP-related ASCVD. We also describe a specific risk pattern for those with CHIP, potentially explaining the enhanced morbidity and mortality associated with cardiogenic shock and ASCVD in this setting.

## Data Availability

All data produced in the present work are contained in the manuscript.

